# *“The ethics approval took 20 months on a trial which was meant to help terminally ill cancer patients. In the end we had to send the funding back”*: a survey of views on human research ethics reviews

**DOI:** 10.1101/2020.07.22.20159533

**Authors:** Anna Mae Scott, Iain Chalmers, Adrian Barnett, Alexandre Stephens, Simon E. Kolstoe, Justin Clark, Paul Glasziou

## Abstract

**Background:** We conducted a survey to identify what types of health/medical research could be exempt from research ethics reviews in Australia.

**Methods:** We surveyed Australian health/medical researchers and Human Research Ethics Committee (HREC) members. The survey asked whether respondents had previously changed or abandoned a project anticipating difficulties obtaining ethics approval, and presented 8 research scenarios, asking whether these scenarios should or should not be exempt from ethics review, and to provide (optional) comments. Qualitative data were analysed thematically; quantitative data in R.

**Results:** We received 514 responses. Forty-three percent of respondents to whom the question applied, reported changing projects in anticipation of obstacles from the ethics review process; 25% reported abandoning projects for this reason. Research scenarios asking professional staff to provide views in their area of expertise were most commonly exempted from ethics review (to prioritise systematic review topics 84%, on software strengths/weaknesses 85%); scenarios involving surplus samples (82%) and N-of-1 (single case) studies (76%) were most commonly required to undergo ethics review. HREC members were 26% more likely than researchers to require ethics review. Need for independent oversight, and low risk, were most frequently cited in support of decisions to require or exempt from ethics review, respectively.

**Conclusions:** Considerable differences exist between researchers and HREC members, about when to exempt from review the research that ultimately serves the interests of patients and the public. It is widely accepted that evaluative research should be used to reduce clinical uncertainties – the same principle should apply to ethics reviews.

## Introduction

Ethics oversight of research is necessary and desirable to protect research participant from excessive physical or psychological risks, or undue burden associated with participation in research. It is increasingly acknowledged that a “one-size-of-ethics-review-fits-all” type of evaluation is inappropriate, and that ethics reviews could be streamlined by developing more risk-proportionate and flexible processes.(1, 2) *Disproportionate* regulation of research – for example, the requirement for low-risk or negligible-risk research to undergo full ethics review – imposes considerable time and cost burdens on researchers and members of research ethics committees.(3, 4) More importantly, it compromises the interests of patients – research hindered or abandoned due to disproportionate regulation can delay the identification of low-value care – that is, care that provides little or no benefit to patients(5) – and adverse and beneficial effects of treatments.(6) In extreme cases, these delays can even be lethal, as shown, for example, by the analysis of the CRASH Trial data, which found that delayed administration of treatment within a trial context (due to fulfillment of consent requirements) increased the patients’ relative risk of death. (7)

Considerable international variation exists in how authorities apply – or do not apply – risk - proportionate review processes. For example, a seven-country trial of a leaflet intended to improve older patients’ involvement in general practitioner consultations was deemed not to require ethics review in 4 countries but required full ethics review in 3 other countries.(8) This “ethics review roulette” is manifested in wide variation of national ethics guidance across different countries.(9) A recent comparison of research ethics review requirements in four countries found that research involving only re-use of publicly available data sets (such as aggregated registry data) is always exempt, but that considerable variation exists for other types of research. For example, surveys of professional staff aimed at improving professional practice were exempt in some but not all countries, and three of the four countries exempt questionnaire or survey research, albeit subject to stipulated conditions such as minimal inconvenience.(10) A recent petition to the Australian government further highlighted the problems faced by Australian researchers when navigating research ethics processes. Many of the over 400 comments prompted by the petition emphasised that “the overburden of bureaucracy in research is harmful to patients” and that a better system “would mean more efficient and better quality outcomes for patients.”(11)

We report here the results of a survey designed to explore the perceptions of Australian health/medical researchers and members of Australian research ethics committees on how the Australian research ethics system might serve the interests of patients more effectively. Specifically, we wished to identify which types of research might be exempted from ethics review because the risks theoretically associated with them are minimal or non-existent.

## Methods

This survey is reported following the Checklist for Reporting Results of Internet E-Surveys (CHERRIES) reporting guideline.(12)

### Sample

Our survey targeted active human health and medical researchers and members of Human Research Ethics Committees (HRECs) in Australia (i.e., it was a “closed survey”, limited only to a known sample rather than open to everyone to complete)

Australian researchers were identified by searching PubMed for publications between 1 April 2019 and 30 June 2019 for which the corresponding author’s affiliation field contained the words “Australia” OR “Australian.” The resulting references (n=4956) were exported to EndNote. The author address field was then searched for the term “.au”. From these, a sample of 900 emails and names was randomly selected (by AB).

A list of 208 Australian HRECs which is maintained by the National Health and Medical Research Council(13) was used to obtain contact information for the HREC’s Administrator (or alternatively Chair or Manager). Where no website was provided, we attempted to identify contact information using web searches. We identified contact information for 175 of the 208 committees in the list.

### Survey instrument

Our survey instrument had 3 parts (see Appendix 1):

- Part 1 (page 1 of the survey): consisted of a demographic question that asked whether the respondent was (i) a human health/medical researcher at an Australian institution; (ii) a former or current member of an Australian human research ethics committee; or (iii) both of the above.
- Part 2 (page 2 of the survey): asked whether the respondent had previously changed or abandoned a research project in anticipation of difficulty in obtaining ethics approval.
- Part 3 (pages 3-11 of the survey): presented eight hypothetical research scenarios and respondents were asked for each whether it should be exempt or not exempt from ethics review in Australia. Respondents were also invited to suggest other research scenarios that could be exempted from ethics review in Australia.

Six of the eight hypothetical research scenarios in Part 3 were derived from an earlier project, comparing the types of research exempt from ethics review in Australia, the United States, the United Kingdom, and The Netherlands(10); two scenarios (Linked Data Sets and N-of-1 (single case) studies) were suggested for inclusion by colleagues who had previously experienced challenges with ethics reviews for those types of research.

We piloted the survey with two colleagues who were otherwise not involved in the project. As the time required for completion proved too burdensome (30 to 40 minutes) we shortened Part 3 of the survey from 8 to 4 research scenarios. However, to still obtain views on all 8 hypothetical research scenarios, we generated four versions of the survey (each containing 4 scenarios) for random distribution among respondents. The resulting survey consisted of 7 pages. The sequence of scenarios was held constant in each survey version – adaptive questioning was not used – but respondents were able to review and change prior answers. (See Appendix 2)

### Survey allocation

The survey was disseminated to active researchers by email. Each email contained a brief introduction to the project and a link to the survey. The survey was hosted on the SurveyMonkey platform. Researchers were randomly allocated one of the four survey versions (A, B, C or D), so the final sample would include a similar number of answers across all the scenarios.

A contact at each HREC was also randomly allocated one of the four survey versions (named H1, H2, H3, H4 – these versions corresponded to researcher survey versions A, B, C, D, respectively. The different naming convention was used to enable separate tracking of the response rates for the HRECs and the researchers). Each HREC contact was asked to distribute the received link to the same survey version (H1, or H2, or H3, or H4) to all their members. This was for logistical reasons, because we did not have the contact emails of the individual committee members.

### Survey dissemination process

The survey was disseminated to researchers in two batches: 295 researchers were contacted on 10 September 2019, and 522 researchers were contacted on 23 September 2019 (each group was sent an email reminder a week later). The survey was disseminated to 175 HREC contacts on 26 September 2019, with an email reminder a week later.

All survey responses were eligible for inclusion if returned before 1 November 2019.

### Analyses

Quantitative data were analysed using R (Version 4.0.0). We estimated the probability of answering that ethics was required using a Bayesian logistic regression model, with fixed effects for respondent type (i.e. researcher only, member of HREC only, both a researcher and HREC member) and random effects for respondents, survey version and scenario. We used vague normal priors for means and Gamma(1,1) priors for precisions. We visually checked the convergence of the chains and compared the estimated probabilities with the observed. The results are presented as the estimated mean probability of answering that ethics is required together with 95% credible intervals. The code to create the quantitative analysis is publicly available (https://github.com/agbarnett/ethics_survey).

Qualitative data were analysed using Excel or NVivo. All of the survey questions (excepting the demographic question) provided respondents with the option to add a comment or further explanation. These were analysed thematically, using an inductive approach.(14) Coding was conducted by two researchers; a third compared the two sets of themes, and rephrased or amalgamated for consistency. Here, we briefly report commonly raised themes underpinning the decisions that a scenario should (or should not) be exempt, together with sample quotes (a fuller discussion of the themes will be reported elsewhere). We corrected minor typographical errors in the quotes but otherwise made no changes.

### Ethics approval and informed consent

[Blinded for peer review] Human Research Ethics Committee provided approval for the project (32214912). The first page of the survey provided information about the survey content and its aims, privacy, data protection, and consent. Participation and completion were voluntary. No incentives for completion were offered.

## Patient and public involvement

Patients and public were not involved in the design or the conduct of this study.

## Results

### Response rates

We received 514 survey responses in total (from both researcher respondents and HREC respondents).

Of the randomly selected sample of 900 researcher emails, 817 successfully received the emails (i.e. the emails did not bounce as erroneous, no longer valid, etc.), and we received 172 responses (response rate 21%).

175 HRECs with available contact email were contacted to disseminate the surveys (versions H1 to H4). We received 342 responses, for an estimated maximum response rate of 24%. (The maximal response rate assumes 175 HRECs of 8 members, which is the minimum number of members on an HREC stipulated by the NHMRC; the response rate may be lower, as HRECs may have more than 8 members). (see Figure 1)

**Figure 1:**
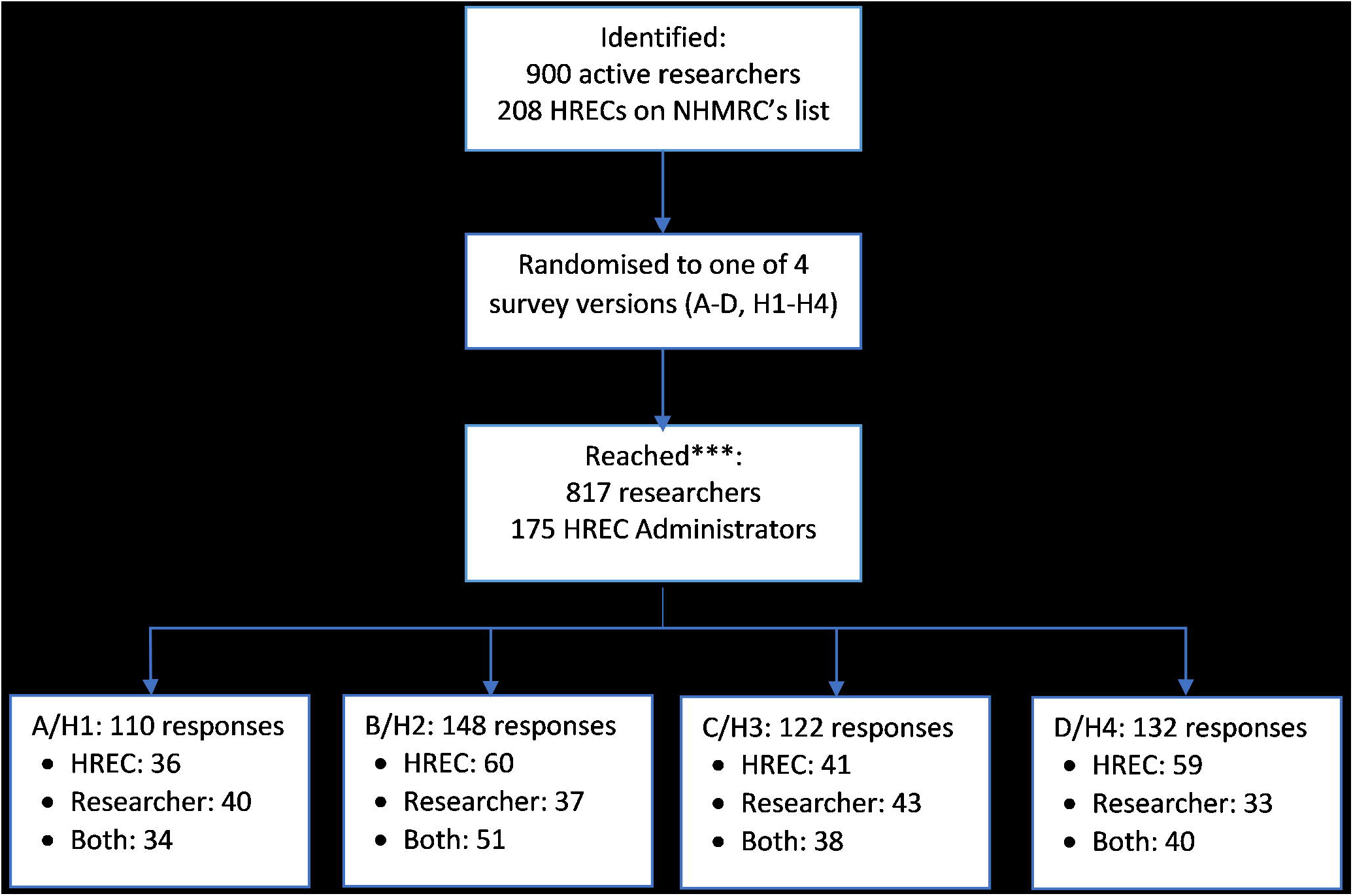
Total number of responses by survey type and respondent type

### Respondents

The median time to complete the survey was 10 minutes (inter-quartile range: 5 to 19 minutes).

Of the 514 responses, 153 were from researchers, 196 were from HREC members, and 163 from individuals who identified themselves as both researchers and HERC members. Two respondents did not identify their status. Responses by survey version and respondent type are summarised in Figure 1.

#### Analysis

#### Changes made to research projects

##### Condensed description of the survey question

I have changed a project from what I had originally envisioned (e.g. changed methods, population) because I was concerned about applying for and obtaining ethics approval (time, complexity, etc.) (for a full description, see Appendix 1)

Of 514 responses to this question: 139 said Yes (27%), 188 said No (37%), 161 stated that this question did not apply to them (31%), and 26 (5%) did not respond.

Of all valid responses (that is, of Yes and No responses only, n=327): 43% said Yes, and 57% said No. (for percentage of both valid responses and all responses to each survey question, see Appendix 4).

53 respondents provided comments explaining the changes they had made.

##### Methods

Respondents reported making changes to outcome measures (e.g. to one considered less invasive), type of study (e.g., to an audit activity with an ethics waiver or to a non-qualitative design), study phase (from phase 2 to phase 1 trial), or recruitment methods (no detail provided). One researcher “*decided to drop a suicide measure and just use a depression measure*.*”* Another *“wanted to change a trial from Phase 1 to Phase 2 before the Ethics Approval Expired*… […] *However, we changed it back to a phase 1 because of the requirements by our ethics would [have] created a great deal of extra work*.*”* Research methods were altered so that a project could be classified as a low/negligible risk rather than higher risk, to avoid a full ethics review. One respondent mentioned that “*If resources and timeframes are tight, I have had to be pragmatic about lowering the risk of the study* (*e*.*g. not asking for identifying information or using secondary data sources with their limitations*).” Another “*altered research methods so that project could be classified as LNR [low/negligible risk] and avoid full ethics review*.*”*

##### Study population

Respondents excluded population groups from studies if they believed that including these groups would elevate the level of study risk or extend the time required for ethics review. Respondents reported “*Avoiding obtaining data from certain groups e*.*g. those under 18 years old, pregnant women, Indigenous peoples, and those with disability” or “[*e*]xcluded analysis of Aboriginal people as [it’s] too hard to get Aboriginal ethics approval” or “chosen a different population to avoid [a] difficult ethics committee and be able to go through a more sensible committee*.*”*

##### Number of centres

Changes were made from multi-site to fewer- or single-site projects to reduce the number of HRECs involved. One respondent stated doing so *“chiefly in the interest of timeliness -- reducing the number of HRECs involved, especially avoiding those known to have few sittings a year and very long lead times, so that projects are viable within the funding period has been critical*.*”* Another mentioned that on *“several occasions [we] conducted single centre projects, where it would have been better to involve willing collaborators at other sites, solely because of the complexity and delay associated with multi-site ethics review for very low risk projects*.*”*

##### Study personnel

Individuals were excluded as study personnel, or a researcher’s role was modified to expedite ethics review. For example, a respondent stated: *“I am an Aboriginal researcher undertaking Aboriginal community health research in my own community. The HREC regarded my situation as a conflict of interest, so I changed my ethics application status from CI-A [Chief Investigator A] to CI-D [Chief Investigator D] so I could continue to engage with my own community*.*”* Another *“did not include research student as investigator with access to identifiable data as timely additional agreements would be required*.*”*

#### Abandoned research projects

##### Condensed description of the survey question

I abandoned a project I had intended to conduct because I was concerned about applying for and obtaining ethics approval (time, complexity, etc.)

Of 514 responses to this question: 81 (16%) of all respondents answered Yes; 242 (47%), answered No; 165 (32%) said the question did not apply to them; and 26 (5%) gave no answer.

Of all valid responses (that is, Yes and No responses only, n=323): 25% of respondents answered Yes, and 75% of respondents answered No.

32 respondents provided information on the types of projects abandoned, or the reasons for abandoning them.

##### Types of projects

Among abandoned studies were those conducted by research students, including a *“Master’s student project – LNR [low/negligible risk] but knew there was no way of getting it through ethics in time*.*”* More generally, another responder commented, student research projects *“often struggle to obtain timely ethics approvals*.*”*

Studies conducted with specific groups were also mentioned, for example, *“prospective studies in patients not able to give consent*.*”* Another commenter noted that *“the ethics approval took 20 months on a trial which was meant to help terminally ill cancer patients. In the end we had to send the funding back*.*”*

##### Reasons provided for abandoning the projects

Burdensome nature of the application processes was cited as reason for abandoning studies – for example, *“Simple case reports and retrospective case series were abandoned because of the time and complexity in applying for ethics approval”*. The burdensome processes were also offered as a more general reason underpinning disinclination to do research *“Current processes for low risk are so cumbersome at my local health service: 100 page submission that still needs to be signed off by CEO of hospital even for low risk with site governance from another committee! Makes me less inclined to do any research with that organisation*.*”*

*“Expense, especially expense in time”* was also offered as a reason for abandoning studies.

Incompatibility with funding timelines was also cited. One respondent mentioned they *“couldn’t secure ethics in time for funding deadline,”* and another *“needed to return a grant due to delays in ethics and governance*.*”* Another respondent mentioned that *“many projects I have been involved with have been abandoned because the ethics approval process takes too long and is too laborious, especially for junior researchers*.*”*

### Scenario 1: N-of-1 (single case) studies in clinical practice

#### Condensed description of the survey scenario

A GP has noticed that some of her patients complain about side effects of a drug. The GP randomises 20 of her patients, to 2 weeks on a placebo and 2 weeks on the medication. The ordering is concealed. All patients consent and keep records of their symptoms. The GP wishes to publish her findings.

Of 213 responses to this question: 51 (24% of valid responses) respondents stated this scenario should <u>not</u> require ethics review, and 162 (76%) respondents stated this scenario <u>should</u> require ethics review (for percentage of both all responses and of valid responses, see Appendix 4). 162 respondents left a comment detailing the reasons for the decision for not requiring (or requiring) ethics review.

#### Reasons for exempting the scenario

The most frequently mentioned reason for exempting this scenario was that it describes standard clinical practice or routine care (25 respondents) – a *“part of necessary treatment/investigations for the patient*.*”* Some respondents (n=6) also noted that *“risks are minimal”* in this scenario.

#### Reasons for requiring ethics review for the scenario

Respondents who required an ethics review for this scenario, emphasised that it requires independent oversight (84 respondents): *“this is research, and requires oversight*.*”* Others required ethics review on the grounds that it needs a review for scientific rigour (n=34) – particularly the need to review the *“design, statistics, to ensure adequate power*.*”* Others (n=18) linked their decision to issues around publishing – one stating that *“The criteria that pushes this over a threshold for ethics review is the desire to publish and disseminate the research further*.*”* Finally, some (n=12) highlighted the privacy issues, stating that as the scenario *“includes private health information of an individual,”* ethics review should be required.

### Scenario 2: No treatment/no behavioural rules imposed

Condensed description of the survey scenario: Study of any design (cohort, randomised control trial, others) that does not require participants to receive any treatment that is not already in routine use, and does not require participants to follow any behavioural rules that are not already applied in routine care.

Of 250 responses to this question: 124 (50%) respondents stated this scenario should <u>not</u> require ethics review, and 126 (50%) respondents stated this scenario <u>should</u> require ethics review. 181 respondents left a comment detailing the reasons for their decision.

#### Reasons for exempting the scenario

Decisions to exempt the scenario frequently emphasised that the scenario represents standard clinical care or routine practice (55 respondents), noting that *“nothing is being done that would not be done anyway, except, basically, counting*.*”* Others referred to the study design (n=32), describing the scenario as a *“clinical audit”* or a *“quality improvement program*.*”* Several also advocated either an altogether exemption or a minimal review (n=4): *“it could be done with either no review or with a rapid, low level review*.*”*

#### Reasons for requiring ethics review for the scenario

Those requiring an ethics review, noted that the scenario requires independent oversight (64 respondents) – *“ethics [review] process provides oversight to ensure that appropriate safeguards are in place*.*”* Many others (n=33) grounded their decision in a need for a review of scientific rigour, highlighting that *“the HREC review also considers the methodology, whether the study has sufficient statistical power to detect an effect etc*.*”* Some respondents (n=20) based their decision on privacy issues, as in the scenario, *“personal medical records are being accessed*.*”*

### Scenario 3: Linked data sets (‘big data’ population health research)

#### Condensed description of the survey scenario

A study involving the use of linked, de-identified administrative data sets (e.g. registries, electronic health records). Data centre holding the original data set assessed the project for privacy issues, feasibility, adequacy of data set creation and analysis. Data sets were held in a secured facility, and individual data could not be seen. Analyses on the linked sets could be performed but not downloaded, nor could the data be downloaded.

Of 192 responses to this question: 126 (66%) respondents stated this scenario should <u>not</u> require ethics review, and 66 (34%) respondents stated this scenario <u>should</u> require ethics review. 119 respondents left a comment detailing the reasons for their decision.

#### Reasons for exempting the scenario

Respondents who thought the scenario should not require ethics review, frequently did so based on the level of risk involved (43 respondents), describing the scenario as posing an *“inconceivable risk”*. Many (n=25) did so on the grounds that ethical considerations had previously been addressed, noting that *“If there were no ethical concerns prior to or throughout the study and there is no identifiable data, there is no reason for further ethical review*.*”* Several (n=4) also suggested that some type of a project review could be appropriate but not a full ethics review, as this type of research *“needs clear accountability, transparency re access but not sure that ethics committee is correct route for this*.*”*

#### Reasons for requiring ethics review for the scenario

Respondents who thought this scenario requires an ethics review most frequently cited the need for independent oversight (21 respondents), as it *“reassures patients whose data is used that an independent body had oversight and considered [t]he risks to them*.*”* Others (n=15) raised privacy and confidentiality concerns, noting that the *“potential for reidentification is relevant*.*”* The ethics review’s role in reviewing projects for scientific rigour was also raised (n=6), as this scenario *“requires evaluation of scientific merit*.*”*

### Scenario 4: Surplus samples or tissues during routine collection in clinical practice

#### Condensed description of the survey scenario

Research may use surplus (extra) tissue or samples obtained from people during routine medical procedures (e.g. extra blood at time of sampling, or tissue not required for diagnosis/testing.

Of 232 responses to this question: 41 (18%) respondents stated this scenario should <u>not</u> require ethics review, and 191 (82%) respondents stated this scenario <u>should</u> require ethics review. 163 respondents left a comment detailing the reasons for the decision.

#### Reasons for exempting the scenario

Those selecting to exempt the scenario from ethics review most frequently mentioned pre-existing consent (8 respondents), as *“Consent to having blood drawn for the subjects should have already covered the potential use for future investigations*.*”* Others (n=5) highlighted that the use of already collected tissues and samples is involved, and therefore this research *“makes good use of waste”*. Several respondents (n=4) also focused on the low potential for increased risk, mentioning that the use poses *“no additional harm*.*”*

#### Reasons for requiring ethics review for the scenario

Respondents most often (53 respondents) justified decisions to require ethics review for this scenario on consent grounds, stating that the patients’ *“tissue belongs to them and must only be used for purposes that they consented to*.*”* Others (n=47) emphasised the need for independent oversight provided by ethics review: *“there should be checks in place to ensure this system is not abused, and that the patient’s interests are protected”*. Review for scientific rigour (n=28) was thought to be needed, for example because the *“circumstances of collection, storage, security, consent and disposal of samples should be documented and approved*.*”* Some comments (n=20) raised the privacy or confidentiality aspects, stating that *“privacy issues need to be assessed before approval*.*”*

### Scenario 5: Quality assurance or audit project

#### Condensed description of the survey scenario

Audit projects are done to produce information about whether care standards comply with national standards and practice guidelines. Those conducting the audit will report the findings to inform planned improvements locally, and they may also disseminate their findings through publication.

*<u>Example A:</u> An audit and feedback project to reduce inappropriate prescribing*.

Of 190 responses to this question: 132 (69%) respondents stated this scenario should <u>not</u> require ethics review, and 58 (31%) respondents stated this scenario <u>should</u> require ethics review.

*<u>Example B:</u> Evaluation of door-to-needle time of patients with acute stroke, to improve treatment initiation*.

Of 189 responses to this question: 124 (66%) respondents stated this scenario should <u>not</u> require ethics review, and 65 (34%) respondents stated this scenario <u>should</u> require ethics review.

*<u>Example C:</u> Direct observation of nursing care for newborns, to identify missed elements of care*.

Of 190 responses to this question: 87 (46%) respondents stated this scenario should <u>not</u> require ethics review, and 103 (54%) respondents stated this scenario <u>should</u> require ethics review.

129 respondents left a comment detailing the reasons for the decision.

#### Reasons for exempting the scenario

Respondents who supported exempting all 3 examples within this scenario, frequently did so based on study design (50 respondents), noting that quality improvement and *“these sorts of audits should not require ethics review*.*”* Others (n=13) based their decision on the low or negligible risk levels involved in the scenario, stating that it involves *“[n]o risk to patients*.*”* The scenario was also considered exempt on privacy/confidentiality grounds (n=11) as *“none of these are taking any personal information*.*”* Some respondents (n=9) also raised the issue of standard practice / routine care, emphasising that *“all [scenarios are] assessing existing standard practice with no impact on those processes*.*”*

#### Reasons for requiring ethics review for the scenario

Those requiring ethics review for all 3 examples within this scenario, were concerned that the scenarios involved direct observation of or interaction with patients (23 respondents), and stated that *“any research involving direct relationship, even just observation, needs to be ethically reviewed*.*”* Others were concerned about publishing issues (n=16), noting that *“journals demand ethics approval*.*”* The need for independent oversight was also provided as a reason (n=8) as *“the participants are in a vulnerable position, either as employees of health services, or due to their status as patients (older, neonate, stroke) etc. Ethics review ensures consent and capacity obligations are met*.*”*

### Scenario 6: Survey/questionnaire of patients, lay persons or carer (non-professional)

#### Condensed description of the survey scenario

A survey or questionnaire of patients, lay persons, or care providers, which does NOT include questions about highly sensitive areas (e.g. suicide, etc.) AND meets one the following criteria:

*<u>Example A:</u> Identity of respondents cannot be readily identified*.

Of 220 responses to this question: 129 (59%) respondents stated this scenario should <u>not</u> require ethics review, and 91 (41%) respondents stated this scenario <u>should</u> require ethics review.

*<u>Example B:</u> Identity of respondents is known only to the research team, but not outside of the team*.

Of 221 responses to this question: 65 (29%) respondents stated this scenario should <u>not</u> require ethics review, and 156 (71%) respondents stated this scenario <u>should</u> require ethics review.

148 respondents left a comment detailing the reasons for the decision.

#### Reasons for exempting the scenario

Respondents supported exempting both examples within this scenario by the low potential for risk (16 respondents), noting that *“risks are negligible*.*”* Others (n=15) mentioned that the study design is *“similar to post marketing / satisfaction surveys”* and there is *“no intervention*.*”* Decision to exempt was often grounded in privacy/confidentiality considerations (n=12), as *“privacy of participants is protected”* or in the content of the questionnaires (n=12), as the *“topics are not highly sensitive*.*”*

#### Reasons for requiring ethics review for the scenario

Comments supporting the requirement to undergo ethics review in both examples most often focused on privacy/confidentiality issues (46 respondents), emphasising that *“just being anonymous may not mean that people cannot be identified*.*”* Those highlighting the need for independent oversight (n=42) mentioned that *“independent scrutiny in an ethical framework ensures protection of participants and their data*.*”* Respondents also pointed out the need for a review of scientific rigour (n=13), noting that *“HREC can help determine if proper care was taken in designing the study*.*”*

### Scenario 7: Interview with patients, lay persons or carers (non-professional)

#### Condensed description of the survey scenario

An interview (one-on-one or in a group), where consent has been obtained. The interview does NOT include questions about highly sensitive areas (e.g. suicide, etc.) AND meets one of the following criteria.

*<u>Example A:</u> Interviewee’s identity is known to the research team but not to the interviewer*. Of 195 responses to this question: 63 (32%) respondents stated this scenario should <u>not</u> require ethics review, and 132 (68%) respondents stated this scenario <u>should</u> require ethics review.

*<u>Example B:</u> Interviewee’s identity is known to the research team and the interviewer, but no one else*. Of 195 responses to this question: 47 (24%) respondents stated this scenario should <u>not</u> require ethics review, and 148 (76%) respondents stated this scenario <u>should</u> require ethics review.

141 respondents left a comment giving the reasons for the decision.

#### Reasons for exempting the scenario

Respondents exempted both examples in this scenario mainly on the grounds of risk level and consent considerations. Respondents who based their decision on the level of risk (20 respondents), stated that *“the administrative burden is not justified by the level of risk*.*”* Respondents focused on the issues around consent (n=6) generally noted that *“If the individuals are participating in the interview / FG [focus group] it implies that consent is present*.*”*

#### Reasons for requiring ethics review for the scenario

Respondents who required both examples within this scenario to undergo ethics review, often highlighted the need for independent oversight of this type of research (71 respondents): *“external oversight for proposed research should be sought [as researchers] may not see potential issues with their research*.*”* Some raised privacy/ confidentiality concerns (n=15), stating that *“someone who understands the issues needs to review the arrangements to protect privacy*.*”* Several (n=7) required the scenario to undergo review but other than an HREC review: one stated that they *“come down in favour of a rapid, low-level review, but a review nonetheless*.*”*

### Scenario 8: Professional staff providing opinion/views in their area of expertise

#### Condensed description of the survey scenario

Professional staff (e.g. hospital, researchers) are asked as volunteers to share their views on their area of expertise, via a face to face interview or a written/online survey.

*<u>Example A:</u> Survey of general practitioners to prioritise systematic review topics*.

Of 226 responses to this question: 190 (84%) respondents stated this scenario should <u>not</u> require ethics review, and 36 (16%) respondents stated this scenario <u>should</u> require ethics review.

*<u>Example B:</u> A survey of systematic reviewers about systematic review software*.

Of 227 responses to this question: 193 (85%) respondents stated this scenario should <u>not</u> require ethics review, and 34 (15%) respondents stated this scenario <u>should</u> require ethics review.

*<u>Example C:</u> Interview with hospital physiotherapists about their experiences in implementing a specific therapy*.

Of 228 responses to this question: 91 (40%) respondents stated this scenario should <u>not</u> require ethics review, and 137 (60%) respondents stated this scenario <u>should</u> require ethics review.

144 respondents left a comment detailing the reasons for the decision.

#### Reasons for exempting the scenario

Respondents who supported exempting all 3 examples within this scenario did so most frequently because of low risk (37 respondents), noting that *“none of these scenarios pose any risk of harms to anyone*.*”* Others focused on study design (n=36), pointing out that these types of studies *“are an essential part of quality improvement and enhance clinical practice*.*”* Respondents also grounded their decision in privacy/confidentiality considerations (n=17) stating that *“individual practitioner privacy [is] maintained*.*”*

#### Reasons for requiring ethics review for the scenario

Respondents who supported ethics review for all three examples, were frequently concerned about privacy/ confidentiality issues (43 respondents), noting the involvement of *“sensitive information regarding individuals*.*”* Others (n=35) highlighted the need for independent oversight as *“staff may be more likely to participate if there has been an independent review*.*”* Respondents were also concerned about publishing issues (n=9), mentioning that *“the presumption is that Ethics approval is needed when the results will be published*.*”*

### Summary plot of scenario responses

None of the presented scenarios were universally exempted from a requirement to undergo ethics review (see Figure 2). Professional staff providing opinion on the areas of their expertise or scope of practice were most often exempted (84– 85% of responses, depending on the example); less frequently exempted were the quality assurance/audit activities (66–69% of responses, depending on the example). (See Appendix 4).

**Figure 2:**
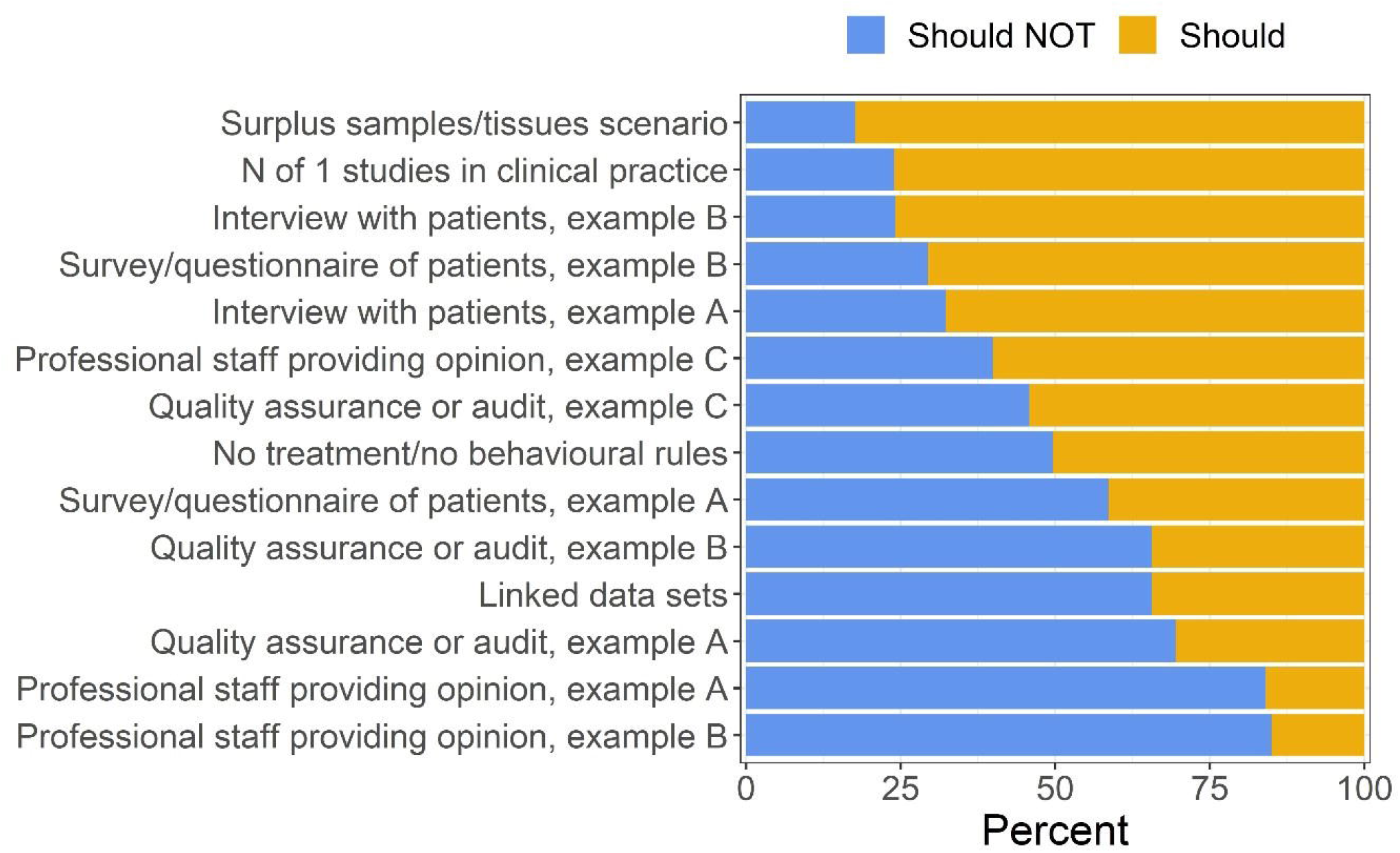
Summary of responses to each scenario

Similarly, no scenario was universally required to undergo ethics reviews. Most commonly, respondents required studies of surplus samples and tissues (82% of responses) and “N of 1” studies in clinical practice (76% of responses) to undergo ethics reviews

### Probability of answering that a scenario requires ethics review by respondent type

Probability of answering that an ethics review is required was the highest for respondents who were members of HRECs; those who were both HREC members and researchers were slightly less likely to require ethics review, but generally, the probabilities are nearly identical for these two groups. Researchers were less likely than both HREC members, and respondents who were both HREC members and researchers, to require an ethics review (Figure 3).

**Figure 3:**
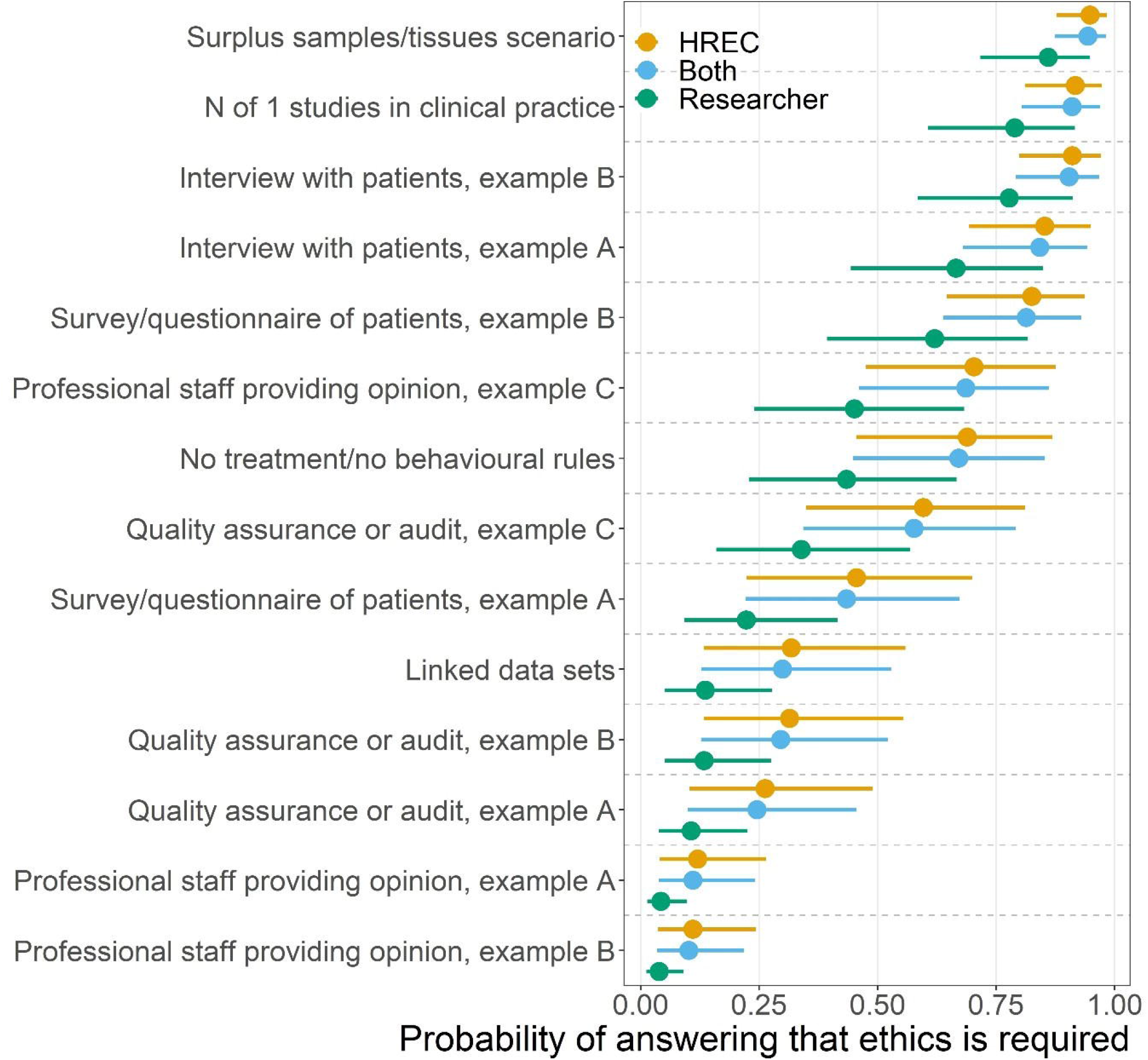
Probability of answering that a scenario requires ethics review by respondent type. Mean estimates (dots) and 95% credible intervals (horizontal lines) from the Bayesian logistic regression model with a fixed effect for respondent type and random effect for scenario.

Overall, members of HRECs were 26% more likely to answer that ethics review was required than researchers, whilst the difference between those respondents who were members of HRECs and those who identified as both researchers and HREC members was 2% (indicating no strong difference). (See Appendix 3).

## Discussion

This survey of researchers and members of research ethics committees in Australia revealed no unanimity about the circumstances in which research ethics review was needed. Based on exemptions listed in guidance in other countries,(10) we identified two particular scenarios (interviews of professional staff about systematic review topics, and evaluation of systematic review software) – for which we found that 84% and 85%, respectively, agreed that ethics review was not required. Conversely, there was no scenario for which over 80% agreed that ethics review was required. Thus, for most scenarios (6 out of 8) there was no clear consensus on whether ethics review should be required. Views differed by the characteristics of the respondents: members of research ethics committees, and those who are both members of HRECs and researchers, were more likely than researchers who were not HREC members to indicate that ethics review was required for all 8 scenarios.

A second important finding was on the impact of seeking ethics approval on research design and planning. Among respondents to whom this question applied, 43% reported changing their research plans because they anticipated obstacles resulting from the ethics review process, and 25% of decided to abandon research projects because of this. A well-functioning ethics review process should be stopping some types of particularly risky research, so some of these decisions to change or abandon projects may have been appropriate. However, the comments also showed that researchers are changing the research they do because of the ethics review process, rather than for sound ethical reasons. For example, proposed low risk student projects were abandoned because they could not be completed within a reasonable timeframe due to uncertainties and delays resulting from ethics review.

Our survey had several limitations. Firstly, it was difficult to identify active researchers. As a proxy, we based our sample on researchers who had published recently. The response rate of around 20% was modest, but the high completion rates and large numbers of comments were testimony to the interest of these respondents. As with much survey research, respondents might have been more interested in the issues of ethics review (or had recent bad experiences), potentially limiting the applicability of our findings more widely. However, generalisability has been improved by using a random sampling approach rather than a snowball recruitment process. Secondly, there was evidence that our survey was forwarded outside our chosen population. This was suggested by 2 respondents identifying themselves as HREC members only responding to the survey versions disseminated to researchers (versions A-D), and 12 researcher responses to surveys disseminated to HREC members (versions H1-H4). However, as the questions and scenarios were identical, this does not significantly undermine the validity of the findings. Thirdly, some of our results may not apply outside Australia as respondents either overtly, or sub-consciously, tailor their responses to the norms of the system they are used to working within. For instance, in the Netherlands and the UK, research only involving medical/healthcare staff and focused on their professional practice is exempt from formal ethics review, so if this survey were completed in these countries, higher proportions or respondents might state an exemption for this particular scenario. Although repeating similar surveys in other countries might help paint a fuller picture, it is likely that responses would need to be corrected for knowledge of existing practice in their locations/countries.

Our findings are consistent with previous research, which has revealed considerable variability in opinions on the need for ethics review. A UK Delphi study achieved consensus on the level of ethical concern for only 14 of 46 questions. Interestingly there was also poor consensus on whether a project was classified as research or audit, and although most audits were considered to be of low ethics concern, some could be judged as high concern.(15) A three-country study of research ethics chairs presented with three cluster randomised trial scenarios also showed considerable variability.(16) Two of the scenarios – a cluster trial of mass media advertising campaigns to increase colorectal cancer screening and a cluster trial of an educational intervention targeting GPs – were split, with about half wanting full review and the others wanting either expedited or no review, suggesting low risk.

This type of disagreement – both among the members of a single ethics committee and across different ethics committees – may lead to undesirable “ethics roulette.”(6) This might be reduced by providing clear case examples in ethics guidance. When researchers and practitioners are considering how to improve care, research ethics committees whose requirements impede these quality improvement initiatives should be challenged to justify their regulatory demands and the resulting efforts and delays that these will entail.

## Conclusion

Our survey shows considerable differences between the researchers and members of ethics committees, about how best to serve the interests of patients and the public. This difference is reflected in conflicting comments left on our survey:

> *“If you don’t have an ethics process/panel then there is no accountability for behaviour”* – respondent who self-identified as a member of HREC only
>
> *“It’s time for this benighted country to catch up and begin to rein in these out-of-control ethics committees”* – respondent who self-identified as a researcher only

Ethics reviews endeavour to protect patient interests, but as we have shown previously, ethics review also has the capacity to do more harm than good.(6, 7) Of particular concern is the potential harm resulting from inhibiting or altering research that could benefit patients, such as quality improvement studies or interviews with professional staff.

When there are uncertainties about which among alternative *treatments* can be expected to do more good than harm, the interests of patients are best served by evaluative research to reduce or resolve the uncertainties. The same general principle, of conducting evaluative research to reduce or resolve the uncertainties, should be applied to ethics review.

## Data Availability

Deidentified data will be available upon reasonable request from the corresponding author

## Acknowledgements

We would like to acknowledge and thank the anonymous peer reviewers for the Journal, for their valuable feedback and suggestions.

