## Appendices for "*“The ethics approval took 20 months on a trial which was meant to help terminally ill cancer patients. In the end we had to send the funding back”*: a survey of views on human research ethics reviews"

### Appendix 1 – Full survey

**PART 1: ABOUT YOU**

1. Which of these options best describes you?

- Member of an Australian Human Research Ethics Committee (former or current member)
- Human health/medical researcher (former or current) working at an Australian institution (e.g. a university, local health district, research institute, etc.)
- Both

**PART 2: CHALLENGES IN THE CURRENT RESEARCH ETHICS ENVIRONMENT**

We would like to ask you how the existing Australian research ethics processes and requirements impact your research.

1. **I have changed a project from what I had originally envisioned** to a different project (e.g. to use different methods, study population) because I was concerned about applying for and obtaining ethics approval (e.g., length of time required to obtain approval, complexity, expense, incompatibility with funder timelines, etc.)

(Please note: we are NOT referring here to modifications made *in response* to requests from the ethics committee, but rather, modifications made *prior to the project’s submission to an ethics* *committee* for review)

- Yes
- No
- This question does not apply to me

**Optional:** If you would like to provide more details on what type of project you **changed**, how you changed it, and/or reasons why, please do so in the space below.

2. **I abandoned a project I had intended to conduct** because I was concerned about applying for and obtaining ethics approval (e.g., length of time required to obtain approval, complexity, expense, incompatibility with funder timelines, etc.)

- Yes
- No
- This question does not apply to me

**Optional:** If you would like to provide more details on what type of intended project you **did not** conduct, and/or reasons why, please do so in the space below.

**PART 3: HYPOTHETICAL SCENARIOS**

We would be grateful to know your views about possible exemption from a requirement to undergo ethics review in Australia, for the scenarios illustrated below.

**Scenario 1: “N of 1” (single case) studies as part of regular, clinical practice**

A patient recently started on a statin has complained to his general practitioner (GP) about muscle pains and fatigue, and says that he is considering stopping the drug. The GP wants to be confident that the statins are likely to be the cause of the patient’s symptoms, so – with the patient’s consent – the patient is allocated to 2 weeks on a placebo and 2 weeks on a statin, but with the ordering of the drug and placebo concealed. Altogether, 20 of the GP’s patients taking a statin and complaining about the unpleasant symptoms, consent to using this approach before deciding whether to stop the statins. The patients keep records of their symptoms throughout the duration of the administration of the drug or placebo. The GP finds that most patients experience unpleasant symptoms whilst taking statins, but the symptoms disappear whilst taking placebo. The GP wishes to use these findings to help her patients and share them more widely through publication.

This type of scenario:

- Should NOT require ethics review
- SHOULD require ethics review

My reasons for this view:

**Scenario 2: No treatment/no behavioural rules imposed**

Study of any design (cohort, randomised control trial, other designs) that does not require participants to receive any treatment that is not already in routine use, and does not require participants to follow any behavioural rules that are not already applied in routine care.

For example, consider a study involving hospitalised patients. The hospital already uses two types of mattress, with some beds using mattress A, and other beds using mattress B. There is uncertainty about whether one of the mattresses is better at preventing bed sores than the other. To address this uncertainty, patients will be randomly allocated either to mattress A or to mattress B, and the incidence of bed sores will be compared in the two groups.

This type of scenario:

- Should NOT require ethics review
- SHOULD require ethics review

My reasons for this view:

**Scenario 3: Linked data sets (‘big data’ population health research)**

A study involving the use of linked, de-identified administrative data sets (e.g. registries, administrative data, electronic health records).

For example, consider a study of the safety of the cardiovascular safety of non-steroidal anti-inflammatory drugs (NSAIDs e.g. Naprosyn, Nurofen). This study involved a de-identified (no names, full date of birth, postcode) data set of people over 75 years of age who used, or did not use, NSAIDs. The prescription records were linked to hospitalisation diagnoses and deaths (with causes) and the rates of fatal and non-fatal heart attacks were compared in users and non-users of the drugs.

The data centre holding the original data set has assessed the proposed project and analyses for privacy impact, feasibility of the study with the available data, and adequacy of data set creation and analysis plans, and has approved the project. The de-identified linked data were held in a secure facility. Individual level data could not be seen on screen. Data were located in a secure environment that allowed analysis but did NOT allow download of the data and did NOT allow download of the analyses. The analyses were checked by data-centre staff for privacy risk before release to the researchers.

This type of scenario:

- Should NOT require ethics review
- SHOULD require ethics review

My reasons for this view:

**Scenario 4: Surplus samples or tissues during routine collection in clinical practice**

Research may use surplus (extra) tissue or samples obtained from people during routine medical procedures. For example, extra blood could be drawn at the time of medically required or routine sampling. Tissue not required for diagnosis or testing could be collected during routine clinical procedures requiring tissue sampling.

This type of scenario:

- Should NOT require ethics review
- SHOULD require ethics review

My reasons for this view:

**Scenario 5: Quality assurance or audit project**

To promote good clinical care, audit projects are done to produce information about whether care standards comply with national standards and practice guidelines. Those conducting the audit will report the findings to inform planned improvements locally, and they may also disseminate their findings through publication in a journal.

For example:

A. An audit and feedback project to reduce inappropriate prescribing, e.g., a chart audit of sedative-hypnotics prescribed to older hospitalised patients.

This type of scenario:

- Should NOT require ethics review
- SHOULD require ethics review

B. Evaluation, pre- and post-simulation training, of door-to-needle time of patients with acute stroke, with the aim of improving treatment initiation.

This type of scenario:

- Should NOT require ethics review
- SHOULD require ethics review

C. Use of direct observation to examine nursing care for sick newborns, to identify missed elements of care, such as phototherapy sessions.

This type of scenario:

- Should NOT require ethics review
- SHOULD require ethics review

My reasons for this view:

**Scenario 6: Survey/questionnaire of patients, lay persons or carer (non-professional)**

A survey or questionnaire of patients, lay persons, or care providers, which does NOT include questions about highly sensitive areas (e.g. suicide, mental illness, HIV status, recreational drug use), AND meets one the following criteria:

1) identity of respondents cannot be readily identified (i.e. the survey or questionnaire is anonymous)

This type of scenario:

- Should NOT require ethics review
- SHOULD require ethics review

2) identity of respondents is known only to the research team and is not disclosed outside of the research team

This type of scenario:

- Should NOT require ethics review
- SHOULD require ethics review

My reasons for this view:

**Scenario 7: Interview with patients, lay persons or carers (non-professional)**

An interview (one-on-one or in a group, such as a focus group) of patients, lay persons, or care providers. Consent has been obtained from participants by the research team. The interview does NOT include questions about highly sensitive areas (e.g. suicide, mental illness, HIV status, recreational drug use), AND meets one of the following criteria:

1) identity of interviewed person(s) is known to the research team but not to the interviewer, and identity cannot be readily identified by the interviewer,

This type of scenario:

- Should NOT require ethics review
- SHOULD require ethics review

2) identity of interviewed person(s) is known to the research team including the interviewer, but is not disclosed to anyone else

This type of scenario:

- Should NOT require ethics review
- SHOULD require ethics review

My reasons for this view:

**Scenario 8: Professional staff providing opinion/views in their area of expertise**

Professional staff (e.g. hospital staff, researchers) are asked – as volunteers – about their views on a topic in their areas of expertise or professional practice, through either a face to face interview or written/online survey.

For example:

A. A survey of general practitioners to prioritise a list of systematic review topics for a Cochrane group.

This type of scenario:

- Should NOT require ethics review
- SHOULD require ethics review

B. A survey asking systematic reviewers to report on the strengths and weaknesses of systematic review software.

This type of scenario:

- Should NOT require ethics review
- SHOULD require ethics review

C. An interview of all physiotherapists in a hospital on barriers and facilitators they have experienced in implementing a specific therapy for patients.

This type of scenario:

- Should NOT require ethics review
- SHOULD require ethics review

My reasons for this view:

**Other research types/scenarios that could be exempted – OPTIONAL**

Do you think there are other type(s) of human health/medical research that could ALSO be considered for exemption from a requirement to undergo ethics review in Australia? (Please input in the space below)

Thank you very much for sharing your views with us.

[click here to submit your answers]

### Appendix 2 – Contents of each of the 4 survey versions

All four survey versions contained the following:

- Part 1: A single demographic question: whether the respondents identify as former or current members of ethics committees, researchers, or both
- Part 2: Two questions, querying whether the respondents have previously modified a project due to concerns about obtaining ethics approval (e.g. timeline incompatibility, complexity), and whether the respondents have previously abandoned a project due to concerns about obtaining ethics approval (e.g. timeline incompatibility, complexity). Both questions had additional spaces for providing detail (optional).
- Part 3: 4 of 8 scenarios, varying by survey version – see below:

Version A/H1: Scenario order:

1. Scenario: Linked data sets (‘big data’ population health research)
2. Scenario: Interview with patients, lay persons or carers (non-professional)
3. Scenario: Quality assurance or audit project
4. Scenario: Professional staff providing opinion/views in their area of expertise

Version B/H2: Scenario order:

1. Scenario: Professional staff providing opinion/views in their area of expertise
2. Scenario: No treatment/no behavioural rules imposed
3. Scenario: Survey/questionnaire of patients, lay persons or carer (non-professional)
4. Scenario: Surplus samples or tissues during routine collection in clinical practice

Version C/H3: Scenario order:

1. Scenario: “n of 1” studies as part of regular, clinical practice
2. Scenario: Quality assurance or audit project
3. Scenario: Survey/questionnaire of patients, lay persons or carer (non-professional)
4. Scenario: Linked data sets (‘big data’ population health research)

Version D/H4: Scenario order:

1. Scenario: No treatment/no behavioural rules imposed
2. Scenario: “n of 1” studies as part of regular, clinical practice
3. Scenario: Interview with patients, lay persons or carers (non-professional)
4. Scenario: Surplus samples or tissues during routine collection in clinical practice

- Final question in all versions of the survey (optional to answer): please suggest other research scenarios that could be exempted in Australia (text-box)

### Appendix 3 – Logic regression model

#### Logistic regression model

We used a Bayesian logistic regression model to estimate the probability of respondents answering that ethics is required. The model examined the effects of the following: 8 scenarios, 4 survey versions (A to D), and 3 respondent types (member of HREC, Researcher, Both).

We adjusted for repeated data from the same respondent using a random intercept.

We tried a more complex regression model where the probability of response for each scenario varied by respondent type. However, the deviance information criterion for this model was substantially greater (indicating a worse fit), hence we present the simpler model.

We plotted estimated probabilities for answering that a scenario should require ethics (see Appendix 3). The plot shows the estimated probabilities for researchers and ‘both’ compared with HREC as the reference category.

The estimated difference in probability of answering that ethics was required between HRECs and researchers was 0.26 (95% CI 0.15 to 0.37, p-value < 0.001). So HRECs were much more likely to answer that ethics was required. The difference between HRECs and those identified as both was 0.02 (95% CI –0.08 to 0.12, p-value = 0.68), indicating no strong difference between HRECs and those who are both researchers and HRECs.

Plot of estimated probabilities (with 95% confidence intervals) for survey version:

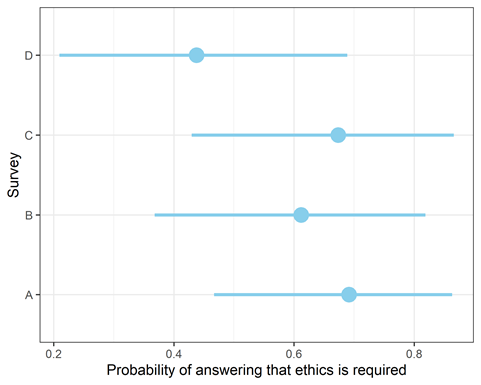

The plot shows the estimated effects of survey version. The estimated difference in probability of answering that ethics was required between survey version A and D was 0.25 (95% CI 0.12 to 0.39, p-value < 0.001). Looking at individual questions in surveys A and D, 77% of researchers answered it should require review to first interview scenario for version A, compared with just 48% of respondents for the same question in version D. The percentages for HRECs were 82% for version A and 59% for version D. Hence there is evidence that responses are varying markedly by survey version.

#### Random effect standard deviations

The table below shows the estimated standard deviations (‘sigma’) for the three random effects from the Bayesian logistic regression model. These were the random effects for: individual respondents, the four survey versions, and the scenarios. The higher the standard deviation, the more variability there was in that group. So there was a much greater variability in responses for individuals and scenarios than for survey versions, but the variation for versions was still relatively large.

| source | sigma |
| --- | --- |
| individual | 1.63 |
| version | 0.755 |
| scenario | 1.65 |

#### Model checking: comparison of observed and predicted probabilities

The plot shows the observed probabilities for (red circle) and the estimates and 95% CIs from the Bayesian model (blue).

The estimates consistently over-estimate the probability for the scenarios with the higher mean probabilities, and this is true for both researchers and HRECs.

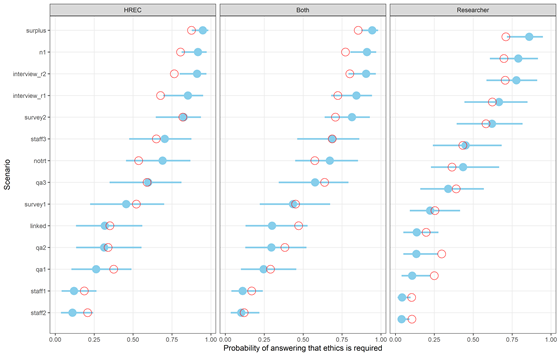

### Appendix 4 – Percent of valid and total responses

The denominator for ‘valid’ responses includes all yes and no answers; it excludes all ‘NA’ answers (those who completed the survey but did not provide an answer for that scenario)

The denominator for ‘total’ responses includes all yes, no, NA, and no response answers.

Table 1: Percent of valid and total responses to ‘modified’ and ‘abandoned’ questions

| **Question: Changes made to research projects** | | | | |
| --- | --- | --- | --- | --- |
|  |  | Freq | % Valid | % Total |
|  | Yes | 139 | 43% | 27% |
|  | No | 188 | 57% | 37% |
|  | <NA or not-responded> | 187 |  | 36% |
|  | Total | 514 | 100 | 100 |
| **Question: Abandoned research projects** | | | | |
|  |  | Freq | % Valid | % Total |
|  | Yes | 81 | 25% | 16% |
|  | No | 242 | 75% | 47% |
|  | <NA or not-responded> | 191 |  | 37% |
|  | Total | 514 | 100 | 100 |

Table 2: Percent of valid and total responses to each scenario

| **Scenario: N-of-1 (single case) studies as part of regular clinical practice** | | | | | | | | | | | | | |
| --- | --- | --- | --- | --- | --- | --- | --- | --- | --- | --- | --- | --- | --- |
|  |  | | Freq | | | | % Valid | | % Total | | | | |
|  | Should NOT require ethics review | | 51 | | | | 24 | | 20 | | | | |
|  | SHOULD require ethics review | | 162 | | | | 76 | | 64 | | | | |
|  | <NA> | | 41 | | | |  | | 16 | | | | |
|  | Total | | 254 | | | | 100 | | 100 | | | | |
| **Scenario: No treatment/no behavioural rules imposed** | | | | | | | | | | | | | |
|  |  | | Freq | | | | % Valid | | % Total | | | | |
|  | Should NOT require ethics review | | 124 | | | | 50 | | 44 | | | | |
|  | SHOULD require ethics review | | 126 | | | | 50 | | 45 | | | | |
|  | <NA> | | 30 | | | |  | | 11 | | | | |
|  | Total | | 280 | | | | 100 | | 100 | | | | |
| **Scenario: Linked data sets (‘big data’ population health research)** | | | | | | | | | | | | | |
|  |  | | | | | Freq | % Valid | | % Total | | | | |
|  | Should NOT require ethics review | | | | | 126 | 66 | | 54 | | | | |
|  | SHOULD require ethics review | | | | | 66 | 34 | | 28 | | | | |
|  | <NA> | | | | | 42 |  | | 18 | | | | |
|  | Total | | | | | 234 | 100 | | 100 | | | | |
| **Scenario: Surplus samples or tissues during routine collection in clinical practice** | | | | | | | | | | | | | |
|  |  | | | | Freq | | | % Valid | | | % Total | | |
|  | Should NOT require ethics review | | | | 41 | | | 18 | | | 15 | | |
|  | SHOULD require ethics review | | | | 191 | | | 82 | | | 68 | | |
|  | <NA> | | | | 48 | | |  | | | 17 | | |
|  | Total | | | | 280 | | | 100 | | | 100 | | |
| **Scenario: Quality assurance or audit project** | | | | | | | | | | | | | |
|  | | “An audit and feedback project to reduce inappropriate prescribing, e.g., a chart audit of sedative-hypnotics prescribed to older hospitalised patients.” | | | | | | | | | | | |
|  | |  | | Freq | | | % Valid | | | % Total | | | |
|  | | Should NOT require ethics review | | 132 | | | 69 | | | 56 | | | |
|  | | SHOULD require ethics review | | 58 | | | 31 | | | 25 | | | |
|  | | <NA> | | 44 | | |  | | | 19 | | | |
|  | | Total | | 234 | | | 100 | | | 100 | | | |
|  | | “Evaluation, pre- and post-simulation training, of door-to-needle time (time between arrival to emergency dept. and administration of treatment) of patients with acute stroke, with the aim of improving treatment initiation.” | | | | | | | | | | | |
|  | |  | | Freq | | | % Valid | | | % Total | | | |
|  | | Should NOT require ethics review | | 124 | | | 66 | | | 53 | | | |
|  | | SHOULD require ethics review | | 65 | | | 34 | | | 28 | | | |
|  | | <NA> | | 45 | | |  | | | 19 | | | |
|  | | Total | | 234 | | | 100 | | | 100 | | | |
|  | | “Use of direct observation to examine nursing care for sick newborns, to identify missed elements of care, such as phototherapy sessions.” | | | | | | | | | | | |
|  | |  | | Freq | | | % Valid | | | % Total | | | |
|  | | Should NOT require ethics review | | 87 | | | 46 | | | 37 | | | |
|  | | SHOULD require ethics review | | 103 | | | 54 | | | 44 | | | |
|  | | <NA> | | 44 | | |  | | | 19 | | | |
|  | | Total | | 234 | | | 100 | | | 100 | | | |
| **Scenario: Survey/questionnaire of patients, lay persons or carer (non-professional)** | | | | | | | | | | | | | |
|  | | “Identity of respondents cannot be readily identified (i.e. the survey or questionnaire is anonymous).” | | | | | | | | | | | |
|  | |  | | Freq | | | % Valid | | | | | % Total | |
|  | | Should NOT require ethics review | | 129 | | | 59 | | | | | 48 | |
|  | | SHOULD require ethics review | | 91 | | | 41 | | | | | 34 | |
|  | | <NA> | | 50 | | |  | | | | | 19 | |
|  | | Total | | 270 | | | 100 | | | | | 100 | |
|  | | “Identity of respondents is known only to the research team and is not disclosed outside of the research team.” | | | | | | | | | | | |
|  | |  | | Freq | | | % Valid | | | | | % Total | |
|  | | Should NOT require ethics review | | 65 | | | 29 | | | | | 24 | |
|  | | SHOULD require ethics review | | 156 | | | 71 | | | | | 58 | |
|  | | <NA> | | 49 | | |  | | | | | 18 | |
|  | | Total | | 270 | | | 100 | | | | | 100 | |
| **Scenario: Interview with patients, lay persons or carers (non-professional)** | | | | | | | | | | | | | |
|  | | “Identity of interviewed person(s) is known to the research team but not to the interviewer, and identity cannot be readily identified by the interviewer.” | | | | | | | | | | | |
|  | |  | | Freq | | | % Valid | | | | | | % Total |
|  | | Should NOT require ethics review | | 63 | | | 32 | | | | | | 26 |
|  | | SHOULD require ethics review | | 132 | | | 68 | | | | | | 54 |
|  | | <NA> | | 49 | | |  | | | | | | 20 |
|  | | Total | | 244 | | | 100 | | | | | | 100 |
|  | | “Identity of interviewed person(s) is known to the research team including the interviewer, but is not disclosed to anyone else.” | | | | | | | | | | | |
|  | |  | | Freq | | | % Valid | | | | | | % Total |
|  | | Should NOT require ethics review | | 47 | | | 24 | | | | | | 19 |
|  | | SHOULD require ethics review | | 148 | | | 76 | | | | | | 61 |
|  | | <NA> | | 49 | | |  | | | | | | 20 |
|  | | Total | | 244 | | | 100 | | | | | | 100 |
| **Scenario: Professional staff providing opinion/views in their area of expertise** | | | | | | | | | | | | | |
|  | | “A survey of general practitioners to prioritise a list of systematic review topics for a Cochrane group.” | | | | | | | | | | | |
|  | |  | | Freq | | | % Valid | | | | | | % Total |
|  | | Should NOT require ethics review | | 190 | | | 84 | | | | | | 73 |
|  | | SHOULD require ethics review | | 36 | | | 16 | | | | | | 14 |
|  | | <NA> | | 34 | | |  | | | | | | 13 |
|  | | Total | | 260 | | | 100 | | | | | | 100 |
|  | | “A survey asking systematic reviewers to report on the strengths and weaknesses of systematic review software.” | | | | | | | | | | | |
|  | |  | | Freq | | | % Valid | | | | | | % Total |
|  | | Should NOT require ethics review | | 193 | | | 85 | | | | | | 74 |
|  | | SHOULD require ethics review | | 34 | | | 15 | | | | | | 13 |
|  | | <NA> | | 33 | | |  | | | | | | 13 |
|  | | Total | | 260 | | | 100 | | | | | | 100 |
|  | | “An interview of all physiotherapists in a hospital on barriers and facilitators they have experienced in implementing a specific therapy for patients.” | | | | | | | | | | | |
|  | |  | | Freq | | | % Valid | | | | | | % Total |
|  | | Should NOT require ethics review | | 91 | | | 40 | | | | | | 35 |
|  | | SHOULD require ethics review | | 137 | | | 60 | | | | | | 53 |
|  | | <NA> | | 32 | | |  | | | | | | 12 |
|  | | Total | | 260 | | | 100 | | | | | | 100 |
